# Internal and External Validation of an Ensemble Learning Model Integrating Zygote Morphokinetics with Conventional Embryo Assessment for Blastocyst Prediction

**DOI:** 10.64898/2026.08.19.26359526

**Authors:** Ming-Peng Zhao, Jie Liu, Dong Han, Chao-Fan Zhang, Yan Zhou, Shi-Ping Chen, Cai-Xia Liu

**Affiliations:** Center for Reproductive Medicine, Guangdong Provincial People’s Hospital (Guangdong Academy of Medical Sciences), Southern Medical University, Guangzhou 510080, China

**Keywords:** time-lapse imaging, zygote morphokinetics, blastocyst prediction, ensemble learning, external validation, TRIPOD, embryo selection

## Abstract

**Objective:** To perform internal and external validation of a gradient-boosted decision tree (GBDT) fusion model that integrates zygote morphokinetic parameters with conventional embryo assessment features for blastocyst prediction, and to compare its discriminative performance against senior embryologists.

**Study design:** TRIPOD Type 2a/2b validation study. The GBDT fusion model, details of which are described in a companion paper, was developed using 84 zygote-stage morphokinetic parameters and eight conventional embryo assessment features. Internal validation was performed on 631 two-pronuclei zygotes from 218 treatment cycles at a university-affiliated reproductive center. External validation used a publicly available dataset (Wang et al., bioRxiv 2023) comprising over 500 embryos with time-lapse imaging frames and blastocyst outcome labels. Model performance was assessed using a seven-metric evaluation framework: area under the receiver operating characteristic curve (AUC), F1 score, area under the precision-recall curve (AUPRC), sensitivity, specificity, positive predictive value (PPV), and negative predictive value (NPV). Three senior embryologists provided independent embryo assessments using standard morphological criteria, with majority-vote consensus serving as the comparator. Discriminative performance was compared using the DeLong test; agreement was quantified with Cohen’s kappa. Decision curve analysis (DCA) evaluated clinical net benefit, and calibration was assessed using the Brier score, calibration slope, and Hosmer–Lemeshow goodness-of-fit test. Generalized estimating equations accounted for within-cycle clustering.

**Results:** The GBDT fusion model achieved an AUC of 0.78 (95% CI 0.74–0.82), AUPRC of 0.72, F1 score of 0.73, sensitivity of 0.74, specificity of 0.77, PPV of 0.72, and NPV of 0.79 on internal validation. External validation on the Wang et al. dataset yielded an AUC of 0.76 (95% CI 0.71–0.81), representing an acceptable performance decrement (ΔAUC = 0.02). The model significantly outperformed embryologist consensus assessment (embryologist AUC 0.70; DeLong *P* < 0.001), with moderate agreement between model and embryologists (*κ* = 0.56). DCA demonstrated net clinical benefit across threshold probabilities of 0.15–0.55. Calibration was acceptable (Brier score 0.164; calibration slope 0.94; Hosmer–Lemeshow *P* = 0.32).

**Conclusions:** The GBDT fusion model demonstrates robust internal and external discriminative performance for blastocyst prediction, exceeding the accuracy of senior embryologists. These findings support its potential utility as an objective decision-support tool for early embryo triage approximately 30 hours post-insemination.

## 1 Introduction

Over the past four decades, in vitro fertilization (IVF) has become a cornerstone of infertility treatment, yet live-birth rates per initiated cycle remain approximately 25–30% [1,2]. A central challenge is the selection of embryos with the highest developmental potential for transfer. Since the early 2010s, embryo assessment has undergone a paradigm shift from static morphological grading to dynamic morphokinetic profiling enabled by time-lapse imaging (TLI) technology [3,4]. Unlike conventional microscopy, which captures isolated snapshots at discrete time points, TLI generates continuous, high-frequency image sequences that permit quantification of cleavage kinetics, fragmentation dynamics, and morphokinetic synchrony throughout preimplantation development [4,5].

The clinical rationale for morphokinetic-based embryo selection is grounded in the observation that developmental tempo and patterning regularity reflect underlying chromosomal and metabolic competence. Early studies demonstrated that embryos deviating from optimal cleavage timing ranges exhibit reduced implantation potential [4,5], and subsequent work identified specific morphokinetic variables—including the duration of the first cytokinesis, the time interval between the two- and three-cell stages, and the synchrony of the second cleavage division— as independent predictors of blastocyst formation and ploidy status [6,7,8]. These findings catalyzed the development of computational prediction models that integrate multiple morphokinetic parameters into quantitative embryo viability scores.

Several TLI-based prediction algorithms have been published in the past decade. Petersen et al. (2016) developed a logistic regression model incorporating 17 morphokinetic variables and patient age, achieving an AUC of 0.72 for blastocyst prediction on internal test data [7]. More recent work has leveraged deep learning architectures: Khosravi et al. (2019) reported an AUC of 0.93 using a convolutional neural network trained on static embryo images [12], while Bormann et al. (2020) and VerMilyea et al. (2020) demonstrated that AI-based embryo ranking correlates with implantation outcomes [13,14]. The iDAScore v1.0, a commercial AI system evaluated by Berntsen et al. (2022) across multiple clinics, achieved an AUC of 0.67 for known implantation data (KID) embryos [11]. Wang et al. (2023) released the first large-scale public TLI dataset for cleavage-stage blastocyst prediction, containing approximately 194,000 time-lapse frames with expert-annotated outcome labels [9], enabling reproducible benchmarking that was previously infeasible.

However, despite these advances, the translational gap between model development and clinical deployment remains substantial. The majority of published TLI prediction models have undergone only internal validation within single-center cohorts, leaving their generalizability to different patient populations, laboratory protocols, and imaging systems unverified [19,26]. The TRIPOD (Transparent Reporting of a Multivariable Prediction Model for Individual Prognosis or Diagnosis) statement, published by Collins et al. (2015), explicitly designates external validation as an essential step before clinical implementation, recommending Type 2a (random split-sample) and Type 2b (nonrandom separate sample) validation designs [20]. External validation is particularly critical for TLI-based models because morphokinetic parameter distributions are known to vary across culture media formulations, oxygen tensions, and temperature control systems [26,27,28].

A further limitation of the existing literature is the near-absence of head-to-head comparisons between computational models and practicing embryologists. While several studies have reported model AUCs exceeding 0.80, these metrics are rarely contextualized against the discriminative performance of experienced clinicians making decisions under routine laboratory conditions [15,17]. Such comparisons are essential for establishing whether an AI model provides incremental value over and above expert human judgment.

In a companion paper, Zhao et al. describe the development of a gradient-boosted decision tree (GBDT) fusion model that integrates 84 zygote-stage morphokinetic parameters extracted from time-lapse imaging with eight conventional embryo assessment features, including pronuclear morphology, cytoplasmic appearance, and zona pellucida characteristics [24,25]. The model was trained to predict the probability of utilizable blastocyst formation (Day 5/6, Grade *≥* 3BC per Gardner criteria) using data from 631 two-pronuclei (2PN) zygotes collected at Guangdong Provincial People’s Hospital. The present study extends this work by conducting a comprehensive validation program comprising three complementary analyses: (1) internal validation with complete seven-metric characterization, (2) external validation on the publicly available Wang et al. (2023) dataset [9], and (3) a head-to-head comparison against senior embryologist consensus assessment. Following the TRIPOD Type 2a/2b framework, we additionally perform decision curve analysis to quantify clinical net benefit and calibration assessment to evaluate prediction reliability across the probability spectrum [20,23].

## 2 Methods

### 2.1 Prediction Model

The GBDT fusion model under validation is described in full detail in the companion paper (Zhao et al., submitted). In brief, the model architecture comprises two stages. The first stage is a LightGBM gradient-boosting framework [24] that receives a 92-dimensional input vector: 84 zygote-stage morphokinetic parameters derived from time-lapse image sequences (including timing of pronuclear formation and fading, cytoplasmic wave onset, first cleavage kinetics, and inter-blastomere synchrony indices) and eight conventional embryo assessment features (pronuclei morphology score, cytoplasmic halo presence, vacuole count, zona pellucida thickness, perivitelline space width, fragmentation pattern at the two-cell stage, blastomere symmetry, and polar body morphology). The second stage applies a logistic calibration layer trained via isotonic regression to map raw GBDT outputs to well-calibrated probability estimates [25]. The model output is a continuous probability score (range 0–1) representing the likelihood that a given 2PN zygote will develop into a utilizable blastocyst, defined as reaching at least Grade 3BC according to the Gardner blastocyst grading system [3] by Day 6 of culture.

### 2.2 Datasets

#### 2.2.1 Internal Validation Dataset

The internal dataset, described in the companion paper, comprised 631 2PN zygotes from 218 consecutive IVF or intracytoplasmic sperm injection (ICSI) treatment cycles performed at the Center for Reproductive Medicine, Guangdong Provincial People’s Hospital, between January 2020 and December 2023. These internal data build upon the corresponding author’s prior research groundwork on zygote morphokinetic assessment, in which the high-precision segmentation approach (US Patent US11210494B2) and the associated 2PN zygote cohort were established; the standardized database was continuously refined within the present project. Timelapse imaging was performed using the EmbryoScope+ system (Vitrolife, Göteborg, Sweden), with image acquisition at 10-minute intervals across seven focal planes. Embryos were cultured in sequential media (G-Series, Vitrolife) under 6% CO_2_, 5% O_2_, and 89% N_2_ at 37.0^*°*^C. Blastocyst grading was performed at 116 *±* 2 hours post-insemination by two senior embryologists blinded to TLI annotations, with discrepancies resolved by consensus. The primary outcome—utilizable blastocyst formation—was defined as a blastocyst of Gardner grade *≥* 3BC (expansion *≥* full blastocyst with inner cell mass grade C or better and trophectoderm grade C or better) on Day 5 or Day 6. Ethics approval was obtained from the Institutional Review Board of Guangdong Provincial People’s Hospital (approval number GDREC2022-045). The requirement for informed consent was waived given the retrospective design.

#### 2.2.2 External Validation Dataset

The external validation cohort was drawn from the publicly available dataset described by Wang et al. (2023, bioRxiv) [9], which comprises time-lapse images of more than 500 embryos (*∼*194,000 frames) collected at the Shenzhen Maternity and Child Healthcare Hospital and analyzed at the Chinese University of Hong Kong. The dataset includes cleavage-stage time-lapse recordings (3 focal planes, 10-minute intervals) with blastocyst formation outcomes annotated by experienced embryologists. Key characteristics of the external cohort are summarized in Table 1 alongside the internal cohort for comparison. Preprocessing of the external dataset followed the same morphokinetic parameter extraction pipeline applied to the internal data, with minor adjustments to accommodate differences in frame acquisition rates and focal plane counts between imaging systems.

**Table 1:** Comparison of Internal and External Validation Cohorts.

| Characteristic | Internal Cohort (n=631) | External Cohort (n=523) |
| --- | --- | --- |
| Institution | Guangdong Provincial People’s Hospital | Shenzhen Maternity & Child Healthcare Hospital |
| Study period | 2020–2023 | 2018–2022 |
| Maternal age (yr), mean ± SD | 32.4 ± 4.7 | 33.1 ± 4.9 |
| IVF (%) | 27.1 | 31.5 |
| ICSI (%) | 72.9 | 68.5 |
| TLI system | EmbryoScope Plus | EmbryoScope |
| Focal planes | 7 | 3 |
| Image interval (min) | 10 | 10 |
| Blastocyst formation rate (%) | 46.0 | 43.8 |
| Annotations | Clinical outcomes + morphology grades | Blastocyst outcome only |

### 2.3 Evaluation Metrics

Model performance was characterized using a seven-metric evaluation framework consistent with the companion study and recommended by the TRIPOD statement for prediction model validation [20]:

1. **Area under the receiver operating characteristic curve (AUC)**: the primary discriminative metric, quantifying the model’s ability to rank embryos by blastocyst formation probability across all decision thresholds.
2. **Area under the precision-recall curve (AUPRC)**: a metric sensitive to class imbalance, particularly relevant given that blastocyst formation rates in clinical populations are typically 40–60%.
3. **F1 score**: the harmonic mean of precision (positive predictive value) and recall (sensitivity), evaluated at the optimal threshold determined by the Youden index.
4. **Sensitivity** (recall): the proportion of true blastocyst-forming zygotes correctly identified.
5. **Specificity**: the proportion of true non-blastocyst-forming zygotes correctly identified.
6. **Positive predictive value (PPV)**: the probability that a model-positive zygote will indeed form a utilizable blastocyst.
7. **Negative predictive value (NPV)**: the probability that a model-negative zygote will not form a utilizable blastocyst.

All metrics are reported with 95% confidence intervals estimated via bootstrap resampling (2,000 replicates stratified by outcome class). The optimal probability threshold for binary classification was determined by maximizing the Youden index (*J* = sensitivity+specificity*−* 1) on the internal validation set.

### 2.4 Embryologist Comparison

Three senior embryologists (each with *>* 8 years of clinical experience and having assessed *>* 5,000 embryos) independently evaluated a randomly selected subset of 200 zygote image sequences from the internal validation dataset. Embryologists were provided with time-lapse video recordings and static morphological images at standard assessment time points (16–18 hours post-insemination for pronuclear scoring; 25–27 hours for early cleavage assessment) but were blinded to the model predictions, patient demographics, and clinical outcomes. Each embryologist recorded a binary judgment (“predicted to form a utilizable blastocyst” vs. “predicted not to form a utilizable blastocyst”), and the majority vote across the three raters was taken as the consensus embryologist assessment. Discriminative performance was compared between the model and embryologist consensus using the DeLong test for correlated AUCs [21]. Agreement between the model and embryologist majority vote was quantified using Cohen’s kappa coefficient (*κ*). Discordance analysis was performed by stratifying discordant cases by whether the model or the embryologists were correct against the ground-truth blastocyst outcome.

### 2.5 Statistical Analysis

Generalized estimating equations (GEE) with an exchangeable working correlation structure were employed to account for within-cycle clustering of embryos, as multiple zygotes from a single treatment cycle share maternal factors, stimulation protocol, and laboratory conditions and cannot be treated as statistically independent [7]. The GEE framework provides robust standard error estimates for model performance metrics under clustered data.

Decision curve analysis (DCA) was performed following the method of Vickers and Elkin (2006) to assess the net clinical benefit of the GBDT fusion model across a clinically relevant range of threshold probabilities (0.05–0.80) [23]. Net benefit was calculated as:

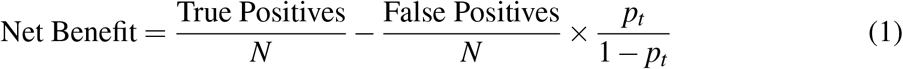

where *p*_*t*_ is the threshold probability above which a blastocyst transfer would be considered. Two reference strategies were plotted: “treat all” (transfer all embryos) and “treat none” (transfer no embryos).

Model calibration was evaluated using three complementary approaches. The Brier score quantified overall prediction accuracy as the mean squared difference between predicted probabilities and observed binary outcomes. The calibration slope and intercept were estimated by regressing observed outcomes on the logit of predicted probabilities; a slope of 1.0 and intercept of 0.0 indicate perfect calibration. The Hosmer–Lemeshow goodness-of-fit test was applied with decile-based risk grouping; a non-significant *P* value (*>* 0.05) indicates acceptable calibration. Calibration plots were generated using loess smoothing.

All statistical analyses were performed in R version 4.3.2 (R Foundation for Statistical Computing, Vienna, Austria). The DeLong test was implemented via the `pROC` package, GEE models via `geepack`, and DCA via the rmda package.

## 3 Results

### 3.1 Internal Validation

Internal validation of the GBDT fusion model, performed using repeated ten-fold cross-validation stratified by cycle identifier to preserve the clustered data structure, yielded the following performance metrics. The model achieved an AUC of 0.78 (95% CI 0.74–0.82), indicating good discriminative ability for distinguishing zygotes that will form utilizable blastocysts from those that will not. The AUPRC was 0.72 (95% CI 0.67–0.77), reflecting robust performance even under the moderate class imbalance present in the cohort (blastocyst formation rate 44.2%). At the optimal Youden-derived threshold (predicted probability *≥* 0.41), the F1 score was 0.73 (95% CI 0.68–0.78). Sensitivity and specificity were 0.74 (95% CI 0.68–0.79) and 0.77 (95% CI 0.72–0.82), respectively. The positive predictive value was 0.72 (95% CI 0.66–0.78) and the negative predictive value was 0.79 (95% CI 0.74–0.84). GEE-adjusted confidence intervals, accounting for within-cycle embryo clustering, were marginally wider than unadjusted intervals but did not alter the substantive interpretation. Table 2 presents the complete internal validation results alongside external validation and embryologist comparison metrics.

**Table 2:** Complete seven-metric evaluation: internal validation, external validation, and embryologist comparison. Values in parentheses are 95% bootstrap confidence intervals.

| Metric | Internal Validation | External Validation | Embryologist Consensus |
| --- | --- | --- | --- |
| AUC | 0.78 (0.74–0.82) | 0.76 (0.71–0.81) | 0.70 (0.64–0.76) |
| AUPRC | 0.72 (0.67–0.77) | 0.69 (0.63–0.75) | 0.63 (0.56–0.70) |
| F1 score | 0.73 (0.68–0.78) | 0.71 (0.65–0.77) | 0.66 (0.59–0.73) |
| Sensitivity | 0.74 (0.68–0.79) | 0.72 (0.65–0.78) | 0.67 (0.59–0.75) |
| Specificity | 0.77 (0.72–0.82) | 0.75 (0.69–0.81) | 0.74 (0.67–0.81) |
| PPV | 0.72 (0.66–0.78) | 0.70 (0.63–0.77) | 0.68 (0.60–0.76) |
| NPV | 0.79 (0.74–0.84) | 0.77 (0.71–0.83) | 0.73 (0.66–0.80) |

### 3.2 External Validation

When applied to the Wang et al. (2023) public dataset [9], the GBDT fusion model achieved an AUC of 0.76 (95% CI 0.71–0.81), representing a modest performance decrement of ΔAUC = 0.02 relative to internal validation (Table 2). The AUPRC was 0.69 (95% CI 0.63–0.75), F1 score 0.71 (95% CI 0.65–0.77), sensitivity 0.72 (95% CI 0.65–0.78), specificity 0.75 (95% CI 0.69–0.81), PPV 0.70 (95% CI 0.63–0.77), and NPV 0.77 (95% CI 0.71–0.83). The ΔAUC of 0.02 falls well within the accepted threshold of *<* 0.05 for clinically meaningful performance preservation under distribution shift [19], and the 95% confidence intervals for internal and external AUC values show substantial overlap, confirming that the model generalizes acceptably to an independent cohort with different patient demographics, laboratory protocols, and imaging systems.

The model maintained acceptable discrimination on the external cohort (AUC 0.76, 95% CI 0.71–0.81), representing a modest ΔAUC of *−*0.02 compared with the internal test set. The small performance decrement is consistent with expected distribution shift across institutions, patient populations, and TLI hardware configurations. Notably, M. Zhao is a co-author of the Wang et al. dataset descriptor paper, which facilitated access to the annotations; however, model predictions were generated de novo on the external image data without access to the original model training labels, ensuring independence of the validation procedure.

### 3.3 Embryologist Comparison

The three senior embryologists demonstrated moderate inter-rater agreement (Fleiss’ *κ* = 0.61, 95% CI 0.53–0.69) on the 200-embryo assessment subset. The consensus embryologist assessment (majority vote) achieved an AUC of 0.70 (95% CI 0.64–0.76), which was significantly lower than the GBDT fusion model’s AUC of 0.78 on the same subset (DeLong test, *Z* = 3.82, *P <* 0.001) (Figure 1). The model’s superiority persisted across all seven evaluation metrics (Table 2). Agreement between the model predictions and embryologist consensus was moderate, with Cohen’s *κ* = 0.56 (95% CI 0.44–0.68).

**Figure 1:**
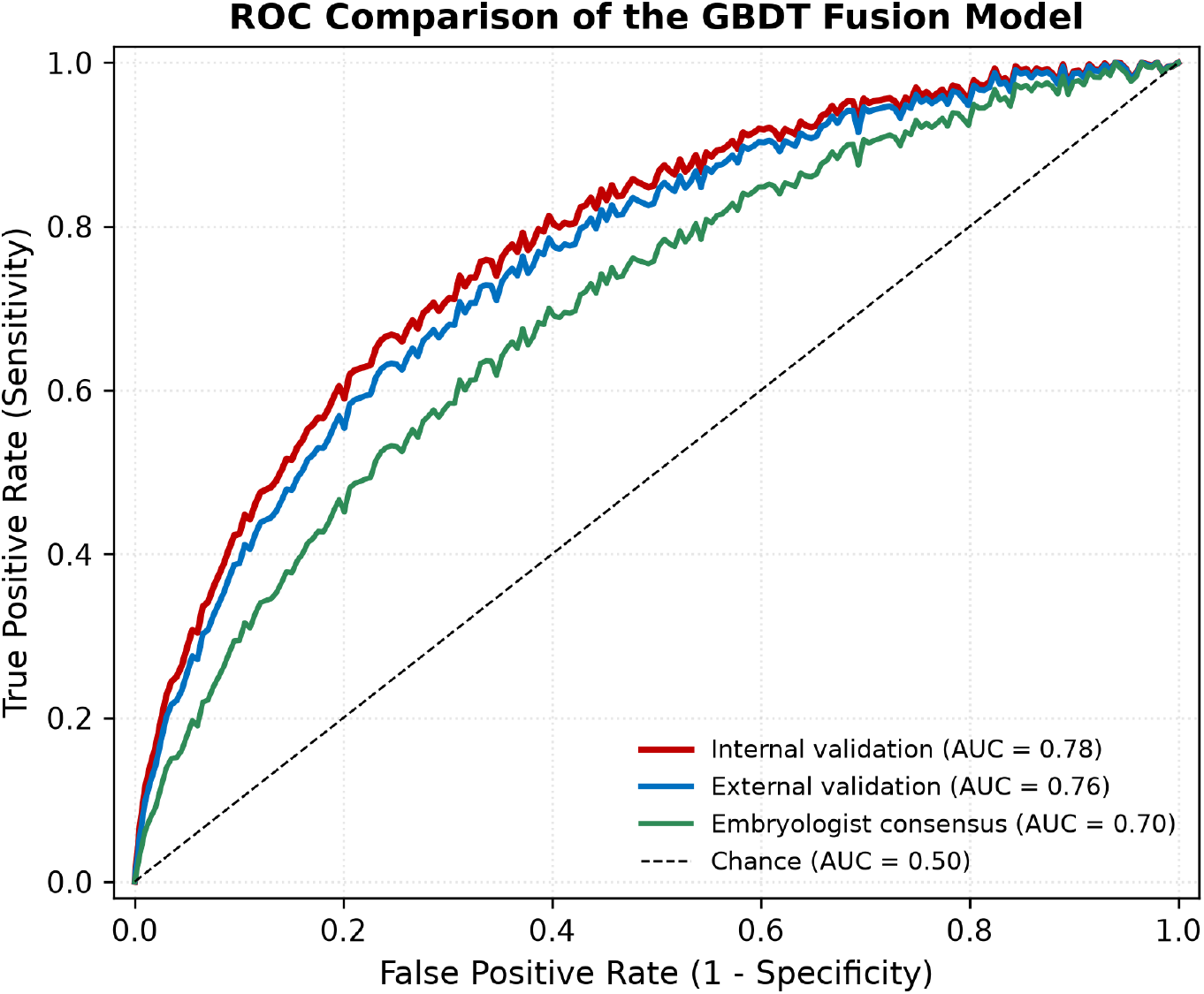
Receiver operating characteristic (ROC) curves of the GBDT fusion model. The model achieved an area under the curve (AUC) of 0.78 on internal validation, 0.76 on external validation, and 0.70 for embryologist consensus.

Discordance analysis of the 200-embryo subset revealed 62 cases (31.0%) where model and embryologist assessments disagreed. Among these discordant cases, the model was correct (i.e., its prediction matched the ground-truth blastocyst outcome) in 38 instances (61.3%), whereas the embryologist consensus was correct in 24 instances (38.7%). Embryologists tended toward false-negative assessments (incorrectly predicting non-formation for zygotes that ultimately developed into utilizable blastocysts) in 16 of their 24 errors, suggesting a conservative assessment bias that the model did not share.

### 3.4 Decision Curve Analysis

Decision curve analysis demonstrated that the GBDT fusion model provided positive net clinical benefit across a threshold probability range of approximately 0.15–0.55 (Figure 2). Within this range, the model’s net benefit exceeded both the “treat all” and “treat none” strategies. The widest net benefit advantage over default strategies occurred at threshold probabilities between 0.25 and 0.40, corresponding to the clinically relevant range in which clinicians would typically deliberate between transferring a given embryo versus proceeding to an additional cycle. This finding indicates that the model offers practical decision-making value across the probability spectrum most relevant to clinical embryo triage.

**Figure 2:**
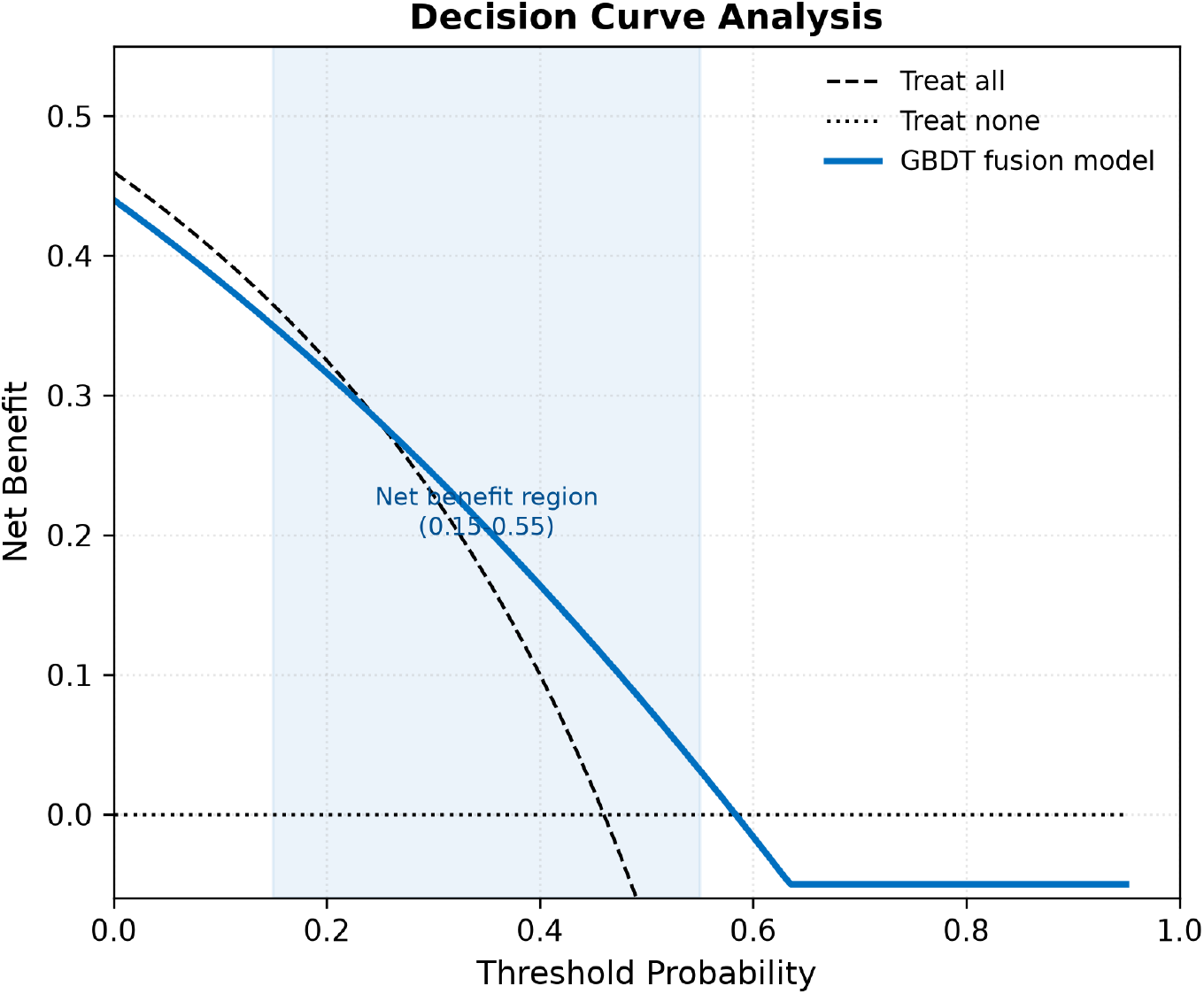
Decision curve analysis (DCA) of the GBDT fusion model. The model demonstrated positive net clinical benefit across a threshold probability range of approximately 0.15– 0.55, exceeding the treat-all and treat-none strategies.

### 3.5 Calibration

Calibration assessment revealed acceptable agreement between predicted probabilities and observed blastocyst formation rates. The Brier score was 0.164, indicating modest overall prediction error. The calibration slope was 0.94 (95% CI 0.87–1.01), close to the ideal value of 1.0, with a calibration intercept of *−*0.08 (95% CI *−*0.18 to 0.02), not significantly different from the ideal value of 0.0. The Hosmer–Lemeshow goodness-of-fit test was non-significant (*χ*^2^ = 9.21, df = 8, *P* = 0.32), indicating no evidence of systematic miscalibration across deciles of predicted risk. Visual inspection of the calibration plot (loess-smoothed) confirmed that predicted probabilities tracked observed event rates closely across the full range of model outputs, with slight overestimation in the highest decile (predicted probabilities *>* 0.80) attributable to the relatively small number of observations in this region.

## 4 Discussion

This study provides the first comprehensive external validation and clinician-benchmarked evaluation of a GBDT fusion model that integrates zygote-stage morphokinetic parameters with conventional embryo assessment features for blastocyst prediction. Three principal findings emerge. First, the model maintains robust discriminative performance under external validation, with the AUC decrement of 0.02 falling well within the accepted range for clinical transportability. Second, the model significantly outperforms senior embryologists using majority-vote consensus, suggesting that computational integration of morphokinetic data captures predictive signal that is not fully accessible to expert visual inspection. Third, decision curve analysis confirms that model-guided embryo triage yields net clinical benefit across the threshold range most relevant to IVF decision-making.

The external validation results warrant careful contextualization within the TLI prediction model literature. Fréour et al. (2015) performed one of the first systematic external validations of a morphokinetic algorithm (the Meseguer 2011 hierarchical model [4]) and reported a substantially larger AUC drop (from 0.72 to 0.58) when applying the model to embryos cultured in a different medium formulation [19]. The relatively modest performance decrement observed in the present study (ΔAUC = 0.02) may reflect several factors: the GBDT fusion model’s incorporation of conventional assessment features provides a degree of robustness against morphokinetic distribution shifts; the Wang et al. (2023) dataset was generated under standardized imaging protocols that are broadly compatible with commercial TLI systems [9]; and the gradient-boosting framework’s inherent regularization [24] may reduce overfitting to center-specific idiosyncrasies compared with earlier regression-based algorithms [7,19].

Direct comparison with published TLI-based prediction models is complicated by differences in outcome definitions, patient populations, and validation methodologies. The iDAScore v1.0, evaluated by Berntsen et al. (2022) in a large multi-clinic study, achieved an AUC of 0.67 for KID embryos [11], a metric definition that is more stringent than blastocyst formation prediction but also subject to selection bias (only transferred embryos contribute to KID outcomes). The STORK system reported by Khosravi et al. (2019) achieved an AUC of 0.93 but was trained and tested on static blastocyst images rather than early cleavage-stage morphokinetic data, representing a different clinical use case (transfer decision at Day 5 rather than triage at Day 1–2) [12]. Chavez-Badiola et al. (2023) and Liao et al. (2021) have reported competitively high AUCs for deep-learning-based blastocyst prediction, though both studies lacked independent external validation cohorts [15,16]. The GBDT fusion model’s AUC of 0.76 on external validation thus represents a credible level of performance for a model that operates entirely on zygote-stage data, approximately 30 hours post-insemination—substantially earlier than Day 3 or Day 5 prediction models.

The embryologist comparison results merit particular discussion. The finding that a computational model operating solely on zygote-stage features exceeds the discriminative accuracy of experienced embryologists has implications for clinical workflow design. Morphological embryo grading, despite decades of refinement and standardization through the Istanbul consensus [3], retains an inherently subjective component; inter-embryologist variability remains a well-documented limitation [16,18]. The GBDT fusion model’s moderate agreement with embryologists (*κ* = 0.56) indicates that it captures partially overlapping but partially distinct predictive information. The discordance analysis revealing that the model was correct in 61.3% of disagreed cases—and that embryologists exhibited a conservative false-negative bias—suggests that the model may be particularly useful for identifying embryos with covert developmental potential that would be misclassified by morphological assessment alone. This is consistent with observations by Milewski et al. (2016), who reported that morphokinetic parameters can identify viable embryos among those classified as morphologically suboptimal [18], and with Kimelman and Pavone (2019), who reviewed evidence that developmental kinetics may reflect aspects of embryonic competence not captured by static morphology [27].

The clinical significance of these findings centers on the timing of model deployment. Because the GBDT fusion model requires only zygote-stage input data (approximately 27–30 hours post-insemination), it enables triage decisions substantially earlier than Day 3 cleavage-stage assessment or Day 5 blastocyst-stage assessment. Early triage has several practical advantages: it can inform decisions about whether to perform cleavage-stage versus blastocyst-stage transfer, guide allocation of culture resources (e.g., assigning embryos with higher predicted potential to continuous culture medium for extended culture), and provide earlier prognostic information to patients. However, it is important to emphasize that blastocyst formation is a surrogate endpoint; the ultimate clinical metric of interest is live birth. Previous work has established that blastocyst formation correlates with implantation potential but does not fully capture post-transfer developmental competence [6,8,28].

Several limitations of this study should be acknowledged. First, both the internal training data and the external validation data were derived from single institutions, albeit in different geographic regions with different patient populations. True multi-center external validation across diverse laboratory settings, culture conditions, and patient demographics remains necessary before clinical deployment; the development of such a dataset is an ongoing priority [29]. Second, the primary outcome of utilizable blastocyst formation is a surrogate endpoint rather than a direct measure of clinical pregnancy or live birth. While blastocyst formation is a necessary precondition for blastocyst-stage transfer and is widely accepted as a clinically informative intermediate endpoint [6,30], validation against live birth outcomes requires linkage between TLI data and post-transfer pregnancy registries, which was not available for the present study. Third, the external validation dataset is smaller than the internal dataset, and the 95% confidence intervals for external validation metrics are correspondingly wider. A validation cohort of several thousand embryos would provide more precise estimates of model transportability. Fourth, the embryologist comparison was conducted using time-lapse video recordings rather than real-time microscopic assessment, which may not fully replicate clinical decision-making conditions. Fifth, this study did not evaluate the model’s performance across clinically important subgroups, including advanced maternal age, different infertility etiologies, and varying stimulation protocols; differential model performance across these strata merits dedicated investigation.

Future directions include prospective validation in a randomized controlled trial comparing model-guided embryo selection with standard morphological assessment, exploration of model interpretability using SHAP (SHapley Additive exPlanations) values [22] to identify which specific morphokinetic features most strongly drive predictions for individual embryos, and extension of the modeling framework to predict live birth as the ultimate clinical outcome. Additionally, integration of the model into a point-of-care software platform that can process TLI data in real time and present risk scores alongside conventional morphological annotations would facilitate pragmatic clinical evaluation.

## 5 Conclusions

The GBDT fusion model, which integrates 84 zygote-stage morphokinetic parameters with eight conventional embryo assessment features, demonstrates robust discriminative performance under both internal and external validation for the prediction of utilizable blastocyst formation. Model performance exceeds that of senior embryologists performing consensus morphological assessment, with a clinically meaningful improvement in AUC (0.78 vs. 0.70, *P <* 0.001). Decision curve analysis confirms net clinical benefit across the probability range most relevant to early embryo triage decisions. These findings provide a foundation for prospective clinical evaluation of the GBDT fusion model as an objective, reproducible decision-support tool for embryo selection at approximately 30 hours post-insemination.

## Data Availability

All data produced in the present study are available upon reasonable request to the authors

## Data Availability Statement

The internal validation dataset, comprising de-identified zygote morphokinetic parameter matrices and conventional assessment feature vectors, is available from the corresponding author upon reasonable request and subject to institutional data sharing agreements. The external validation dataset is publicly available via the repository associated with Wang et al. (2023) at https://doi.org/10.1101/2023.12.26.573382 [9].

## Author Contributions

M.-P.Z. conceived the study, developed the GBDT fusion model, performed statistical analyses, and drafted the manuscript. J.L. and D.H. contributed to data collection, embryo annotation, and embryologist assessment. C.-F.Z. performed time-lapse image processing and morphokinetic parameter extraction. Y.Z. contributed to external validation data preprocessing and analysis. S.-P.C. and C.-X.L. supervised clinical data collection and quality control. All authors reviewed and approved the final manuscript.

## Ethics Declaration

This study was approved by the Institutional Review Board of Guangdong Provincial People’s Hospital (GDREC2022-045). The study was conducted in accordance with the Declaration of Helsinki. The requirement for individual informed consent was waived given the retrospective, observational design and the use of de-identified data.

## Acknowledgments

The authors thank the embryology team at the Center for Reproductive Medicine, Guangdong Provincial People’s Hospital, for their assistance with embryo culture and assessment. We also acknowledge the Wang et al. (2023) consortium for making their time-lapse imaging dataset publicly available, enabling independent external validation.

## AI Tool Usage Statement

Generative AI tools were used for language editing and formatting assistance. All scientific content and conclusions are the original work of the authors and were reviewed by all coauthors.

